# An at-home and electro-free COVID-19 rapid test based on colorimetric RT-LAMP

**DOI:** 10.1101/2022.12.21.22283781

**Authors:** Diem Hong Tran, Hau Thi Tran, Trang Nguyen Minh Pham, Le Minh Bui, Huong Thi Thu Phung

## Abstract

**Purpose:** In the fight against virus-caused pandemics like COVID-19, the use of diagnostic tests based on RT-qPCR is essential but sometimes limited by their dependence on expensive, specialized equipment and skilled personnel. Consequently, an alternative nucleic acid detection technique that gets over these restrictions, called loop-mediated isothermal amplification following reverse transcription (RT-LAMP), has been broadly investigated. Nevertheless, the developed RT-LAMP assays for SARS-CoV-2 detection still require laboratory devices and are electrically dependent, limiting their widespread use as rapid home tests. In this work, a flexible RT-LAMP assay that gets beyond the drawbacks of the available isothermal LAMP-based SARS-CoV-2 detection was developed, establishing a simple and effective at-home diagnosis tool for COVID-19.

**Methods:** A multiplex direct RT-LAMP assay modified from the previously developed test was applied to simultaneously identify the two genes of SARS-CoV-2. We used a colorimetric readout, lyophilized reagents, and benchmarked an electro-free and micropipette-free method that enables sensitive and specific detection of SARS-CoV-2 in home settings.

**Results:** Forty-one nasopharyngeal swab samples were tested using the home-testing RT-LAMP (HT-LAMP) assay developed, showing 100% agreement with the RT-qPCR results.

**Conclusions:** This is the first electrically independent RT-LAMP assay successfully developed for SARS-CoV-2 detection at home setting. Our HT-LAMP assay is thus an important development for diagnosing COVID-19 or any other infectious pandemic on a population scale.

## 1. Introduction

In December 2019, a new coronavirus (SARS-CoV-2) produced an epidemic of COVID-19, a deadly pneumonia disease that quickly spread over the world, resulting in a worldwide outbreak [1]. The causal agent has been identified as a new coronavirus known as SARS-CoV-2 [1]. Since then, hundreds of SARS-CoV-related molecular diagnostic tests have been developed. Specific host antibodies, viral proteins, and viral RNA are the main diagnostic targets, each with its own set of benefits and drawbacks in accurately detecting SARS-CoV-2 during its infectious period in order to more effectively prevent its spread [2]. Professional experience in sampling, performing the reaction, and analyzing the outcomes is required by the most widely used and approved detection method in the world. In addition, it necessitates the use of specialized machines and chemical reagents, as well as advanced sample collection and transportation procedures. When confronted with recent, unprecedented testing volumes, this is especially true.

Diagnostic quantitative real-time PCR following reverse transcription (RT-qPCR) assays for detecting SARS-CoV-2 virus have been critical in preventing and managing the COVID-19 outbreak [3]. RNA purification, reverse transcription, and quantitative PCR are all common procedures for detecting viral RNA in patients [3]. These procedures are time-consuming and necessitate the use of multiple biochemical reagents, low-grade devices, and qualified personnel, and they can be done only at central laboratories. As a result, it is unable to handle the rapidly increasing demand for suspected and silently infected patients. However, with the emergence of asymptomatic infections and their potential for transmission, the number of people who need to be screened is rapidly increasing. Apart from the indicated viral RNA, detection methods based on immunoglobulin (IgM/IgG) antibodies have been expected to identify SARS-CoV-2 rapidly and easily for COVID-19 testing at point-of-care [4,5]. IgM antibodies, on the other hand, are produced between 4 and 10 days after infection, but the IgG response takes about 2 weeks [6]. As a result, low-abundance antibodies in the sample will produce false negative results in the early stages of illness. Therefore, there is a need for diagnostic tools that can detect SARS-CoV-2 RNA precisely and easily-to-use at home for the rapid evaluation of the patients’ treatment impact and prognosis.

Different nucleic acid isothermal amplification approaches for SARS-CoV-2 RNA detection have been developed successfully [7,8]. Among them, colorimetric reverse-transcription loop mediated isothermal amplification (RT-LAMP) appears to be the most attractive alternative diagnostic test because of its increased simplicity, quicker time to achieve a result, and lower cost. Nevertheless, these tests still require basic laboratory equipment and devices to be performed. Consequently, a rapid test based on nucleic acid amplification that can identify the presence of SARS-CoV-2 RNA quickly at home is currently lacking. In this study, we developed a COVID-19 home-testing RT-LAMP (HT-LAMP) kit that is quick, reliable, sensitive, and electro-equipment-free. Our test employs a colorimetric readout to enable reliable SARS-CoV-2 detection while skipping the RNA extraction step. The COVID-19 HT-LAMP kit benefited from freeze-dried reagents and disposable bodywarmers, attempting to make COVID-19 molecular diagnosis more accessible and facilitating large-scale implementation even in settings with limited economic or infrastructural resources.

## 2. Materials and Methods

### 2.1 SARS-CoV-2 Specimen Collection

The SARS-CoV-2 Specimen Collection Kit is designed to collect patient samples specifically for the HT-LAMP assay. Sample Collection tube 1 and Dilution tube 2 contain 400 and 980 µl of nuclease-free water, respectively. Accordingly, nasal or nasopharyngeal samples were collected with a sterile sampling swab, and the swab sample was dipped into Sample Collection tube 1. Gently swirl the swab in the tube, dip the swab up and down in the tube 10 times. Then, an eye dropper with a volume of 200 µl is used to mix the solution in tube 1 and transfer one drop with a volume of approximately 20 µl to the Dilution tube 2. Squeeze and release the dropper gently several times to mix the solution in tube 2. For samples collected in the viral transport medium (VTM) or the universal transfer medium (UTM), mix the sample with an eye dropper and transfer one drop to Tube 2.

### 2.2. HT-LAMP Assay

The HT-LAMP assay was a colorimetric multiplex RT-LAMP reaction using two sets of specific primers to simultaneously detect distinctive sequences of the N and Orf1ab genes of SARS-CoV-2 that were designed and optimized in the previous study [7]. Primers were synthesized by Phu Sa Biochem (Can Tho, Vietnam). A colorimetric multiplex RT-LAMP reaction was carried out as described previously [7]. Briefly, the reaction (20 μl) consists of 10 μl of WarmStart® Colorimetric LAMP 2X Master Mix (DNA & RNA) (NEB, MA, USA), 1.6 µM each FIP/BIP primer, 0.2 µM each F3/B3 primer, and 0.4 µM each LoopF/LoopB primer. The HT-LAMP assay using lyophilized reagents was prepared and performed as in the earlier work [7]. The amplification products were visualized by the color shifting from red to yellow of the testing reaction, which is based on a pH-sensitive indicator as instructed by the manufacturer.

### 2.3. Evaluation of HT-LAMP Kit

Forty-one nasopharyngeal and oropharyngeal swab specimens of suspected COVID-19 patients were utilized to evaluate the clinical performance of the COVID-19 HT-LAMP kit. The specimens were collected in VTM/UTM or nuclease-free water [7]. The samples were kept at -80 °C until analysis. Viral RNA from clinical samples was isolated using the QIAamp RNA mini kit (Qiagen, USA), and the presence of SARS-CoV-2 was confirmed by the Luna® Universal One-Step RT-qPCR Kit using the F3/B3 primer set. The collected specimens were processed as described above, and the diluted samples in tube 2 were transferred by one drop (approximately 20 µl) to the lyophilized reaction tube using an eye dropper with a capacity of 200 µl. The reactions were incubated in the heating pad for 60 minutes, and the results were determined by the reaction color after incubation.

## 3. Results

### 3.1. HT-LAMP Assay Independent of Electrical Equipment

In general, the improvements in the colorimetric RT-LAMP assay made thus far result in more straightforward, reliable, and sensitive SARS-CoV-2 detection tests. However, the assay still needed specialized lab tools, such as temperature-controlled incubators or fine pipettes. As a result, we investigated methods for adapting the RT-LAMP protocol completely to home settings. RT-LAMP reactions require steady incubation temperatures of 60–65 °C due to their isothermal nature. High-end instrumentation and the most basic arrangement, in which boiling and room temperature water are combined in a certain ratio and then kept insulated, can both be used to produce a temperature-controlled reaction environment. Previously, for home-based testing, a commercially available sous-vide heater (water bath) was proposed to be successfully utilized [9]. However, this approach still demands an electrical device and a water-based setup, which are not always available and convenient for in-home testing. With the aim of establishing the assay free of electrical devices, we employed commercially available, inexpensive, convenient, disposable and instrument-free thermopad bodywarmers. Typically, the bodywarmer comes with a special textile adhesive. The heat pad warms up automatically as it comes into contact with oxygen and can provide natural heat for up to 6 or 12 hours.

Thermopads needed to be examined to see if they could serve as thermal incubators by seeing if their operating temperatures stayed largely steady and achieved the ideal range for LAMP reactions, between 60 and 65 °C. Here, the heating capabilities of four different commercial bodywarmers were evaluated. The results revealed that four different types of thermopads took at least 15 minutes to attain their peak temperatures (Figure 1A). The weakest performer was type 3, which had the lowest maximum temperature and lost heat the fastest over time. The type 2 and type 3 could reach temperatures of 55-60°C between 20 and 30 min and sustain the heat for up to 70 min. Type 4 undoubtedly performed the best in terms of maximum temperature reach, stability, and heat maintenance. Multiple tracking data also shows that the type 4 thermopad can provide the heating temperature and duration necessary for the LAMP responses (Figure 1B).

**Figure 1.**
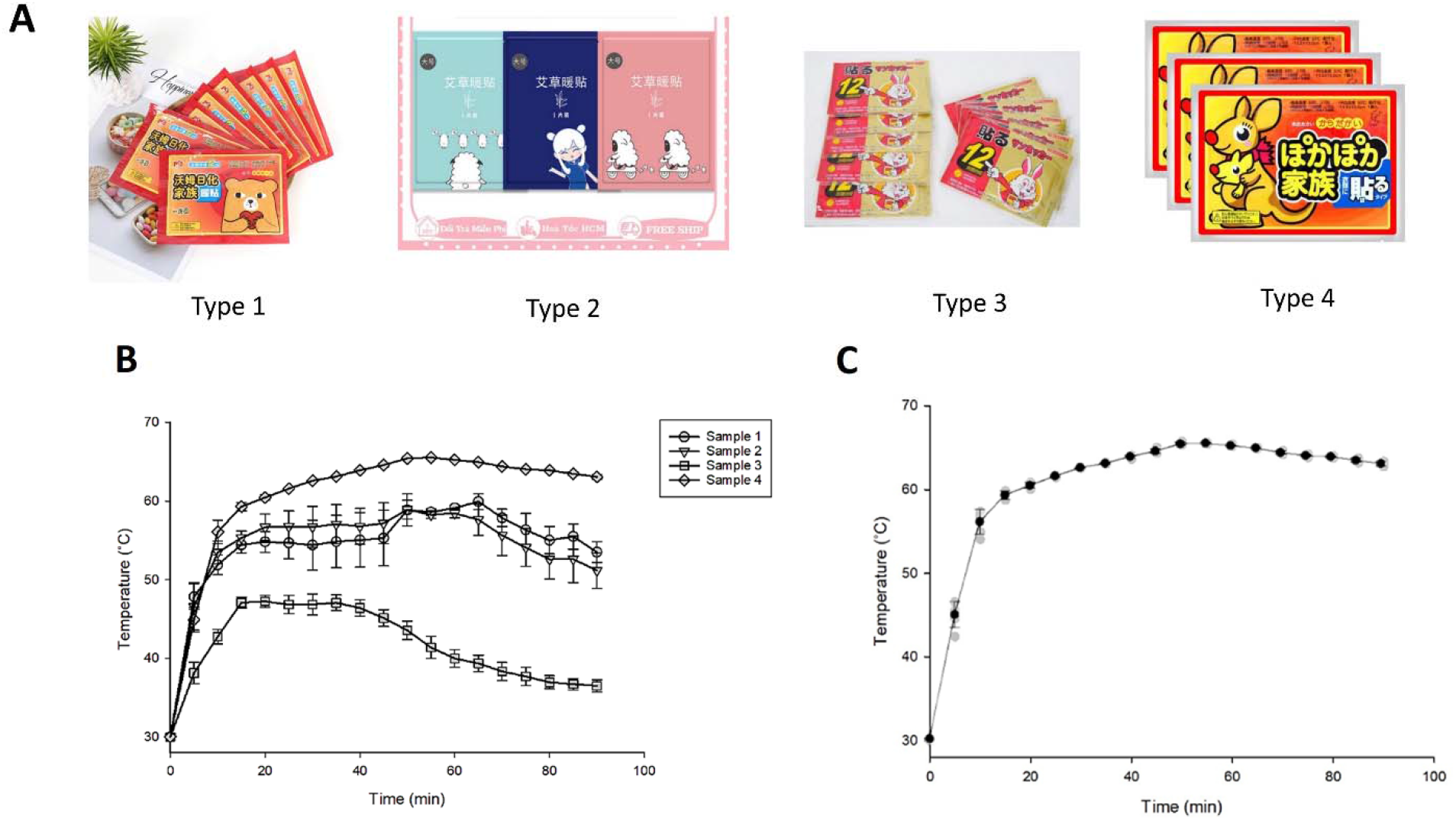
Temperature monitoring of the thermopads. (A) Types of disposable bodywarmers tested. (B) The temperature was recorded every 5 minutes. Using 4 different pads of each type, the experiment was conducted 5 times independently. (C) Following the seal’s removal, the type 4 thermopad’s temperature was measured every 5 minutes. Using 15 different pads, the experiment was conducted 15 times independently. The line represents the average values. Error bars indicate the standard deviation.

When it comes to using pocket bodywarmers, incubation time is one of the critical parameters that must be defined. At the beginning of the heating process, the HT-LAMP reactions were placed on the pads. After 50 min, the reaction tubes showed the expected color shift from pink to yellow, as well as products with better quality signals (Figure 2). Thus, to guarantee the outcome, 60 min were selected to perform the HT-LAMP assay.

**Figure 2.**
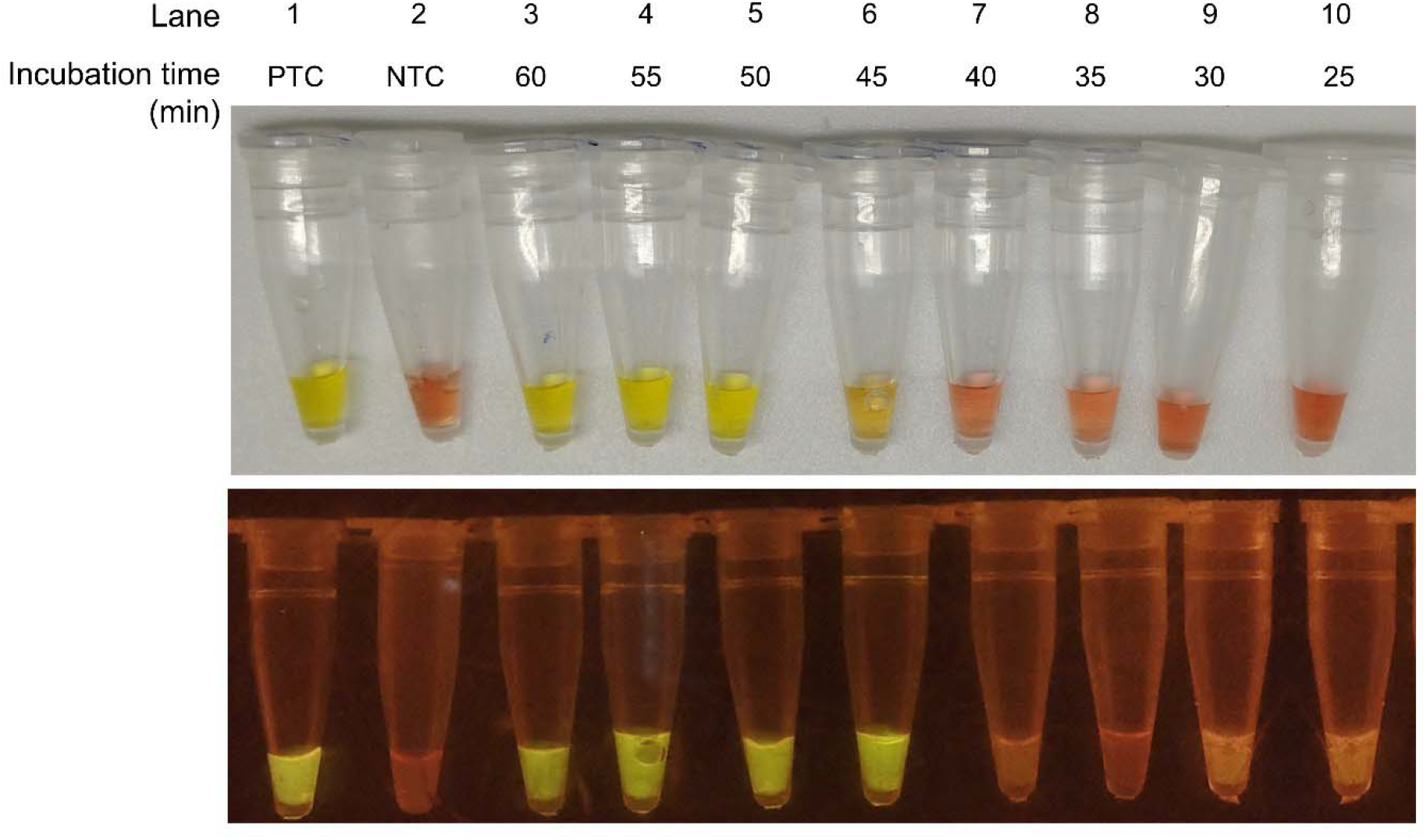
Optimization of the incubation time for HT-LAMP assay. The LAMP reaction containing 10^2^ copies of SARS-CoV-2 genomic-RNA template (7) was incubated in bodywarmers for 25 to 60 min. The color was captured by the personal mobile phone (upper panel) and the SYBR Green I fluorescent signal was visualized by a blue light transilluminator (lower panel). PTC stands for positive-template control; NTC stands for non-template control.

### 3.2. LOD of HT-LAMP Assay

Multiplex RT-LAMP was carried out to identify the LOD of the HT-LAMP assay. Extracted genomic-RNA of SARS-CoV-2 was quantified via a standard curve based on the RT-qPCR Ct-value as described in the previous study [10] and then serially diluted at the indicated concentration. The LOD value was identified as one copy per reaction µl, which is equivalent to 20 copies per reaction (Figure 3). These results indicate a robust performance of the colorimetric HT-LAMP assay across a broad range of genomic-RNA samples, meeting the clinical requirement of the viral load of SARS-CoV-2 in clinical samples [11]. In particular, a study conducted on more than 3303 patients who were confirmed positive for SARS-CoV-2 estimated that the viral load in swab samples was in the range of 10 to 10 copies per mL of sputum [11]. Thus, these viral loads can be easily measured using our HT-LAMP assay.

**Figure 3.**
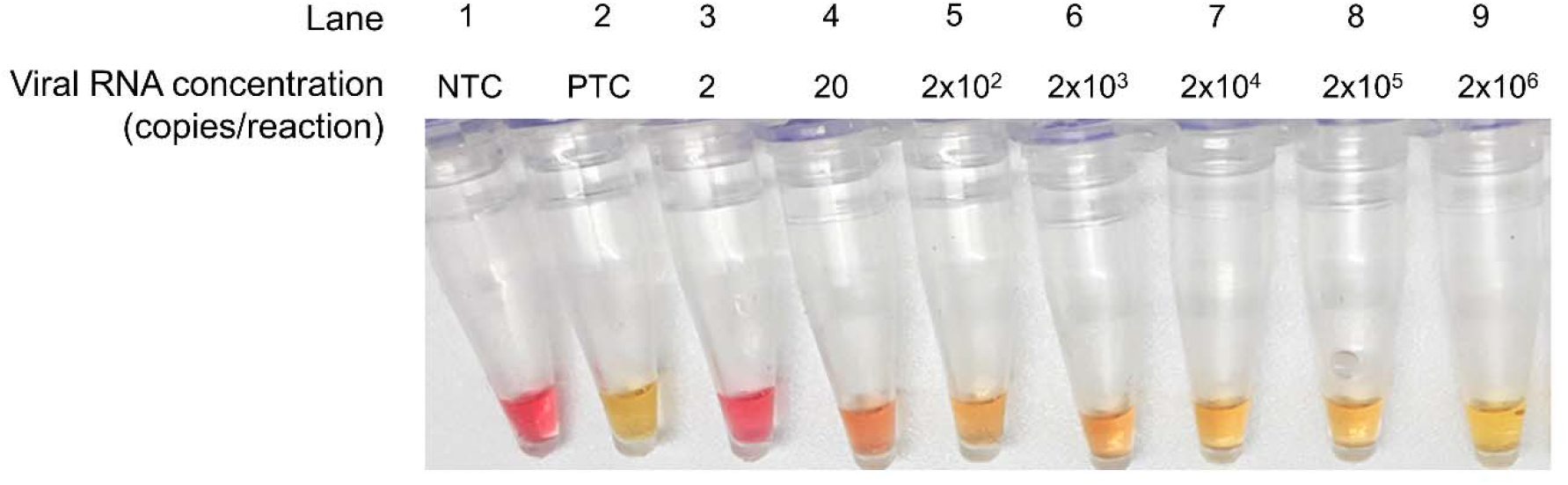
The LOD of HT-LAMP assay. The viral RNA was serially diluted in nuclease-free water to the indicated concentrations, and 20 µl of the diluted sample was added into the HT-LAMP reaction.

### 3.3. HT-LAMP Assay Protocol

The HT-LAMP kit includes a ready-to-use freeze-dried mixture that comprises all the components needed for detection. RNA targets are amplified and detected by the color change of the reaction from red to yellow. The assay includes a non-template sample serving as a negative control (NTC) to confirm the absence of contamination. PTC samples are supplied to verify that the provided reactions are not faulty. A simple eye dropper allowing for controlled dispensing is used instead to transfer a small amount of liquid, eliminating the requirement for an expensive micropipette during the assay in home-based testing.

The protocol of the HT-LAMP assay is illustrated in Figure 4 accordingly. The collected specimens are first processed, as mentioned above. Next, use a new 200 µl dropper to transfer one drop of solution from tube 2 to a lyophilized LAMP reaction tube. Use the dropper to gently squeeze and release to dissolve and mix the solution in the tube well. The solution is now bright pink. At the same time, the bodywarmer is activated. Cap the reaction tube tightly and put it in the center of the heating patch. Fold the patch so that it covers the entire reaction tube. Incubate the reaction for 60 min in the heating pad, then examine the results by color. The two controls are required to accurately exhibit the color as expected. The positive control gives a yellow color, and the negative control keeps the original pink color. The positive sample changes from pink to yellow, while the negative sample remains pink.

**Figure 4.**
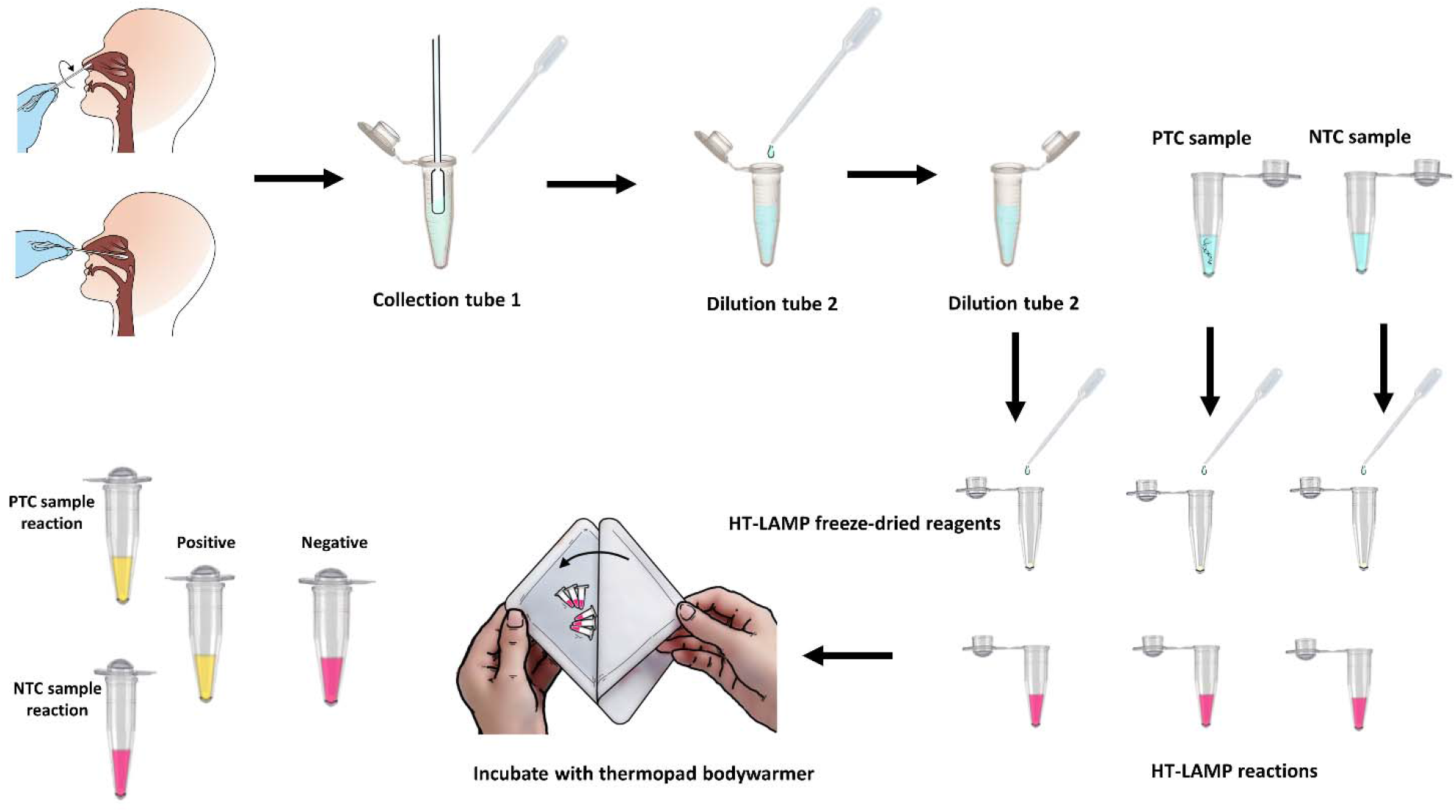
HT-LAMP for SARS-CoV-2 detection in limited and home-settings. A sterile sampling swab is used to collect nasal or nasopharyngeal samples. The swab is then dipped up and down and gently swirled into Sample Collection tube 1 for 10 times. The solution in tube 1 is then mixed by gently squeezing and releasing an eye dropper, and one drop is transferred to Sample Dilution tube 2. To mix the fluid in tube 2, gently squeeze and release the dropper several times. Next, transfer one drop of solution from tube 2 to a lyophilized LAMP reaction tube using a fresh dropper. NTC and PTC samples are similarly transferred into control reaction tubes, respectively, by dropping using fresh and separated droppers. The warmer is prepared by opening the package, removing the warmer, and shaking it a bit. Wait one to two min for the oxygen activation. Remove the protective foil and place the reaction tubes in the center of the adhesive part of the patch. Fold the patch in half carefully to completely cover the tubes. Wait for 60 min, then open the patch and examine the results by the color shift of the reactions. After a single usage, discard the bodywarmers as general garbage.

### 3.4. Evaluation of HT-LAMP

Nasopharyngeal swabs were collected from 41 suspected COVID-19 patients. The gold-standard RT-PCR results showed that 21 samples were positive and 20 samples were negative. To validate the home-testing kit performance, all 41 samples were tested using the HT-LAMP assay. The results turned out to be that the HT-LAMP assay had 100% sensitivity and specificity when compared to RT-PCR. The findings indicate that the HT-LAMP assay can be used as an alternative tool for SARS-CoV-2 detection, allowing for a rapid and electro-equipment-free diagnosis in a home setting.

**Table 1.**
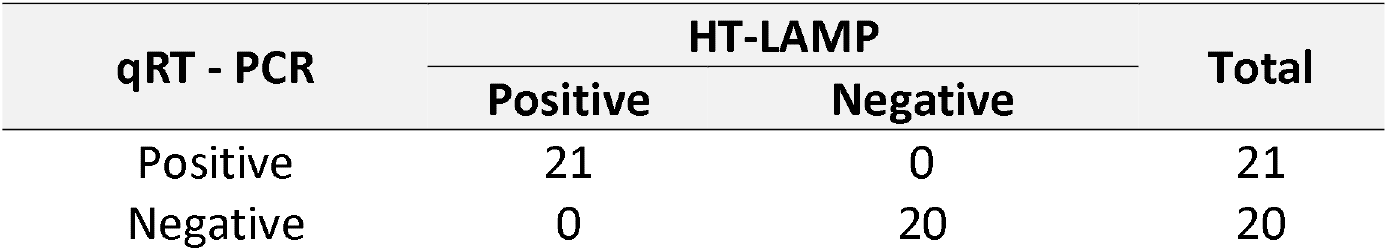
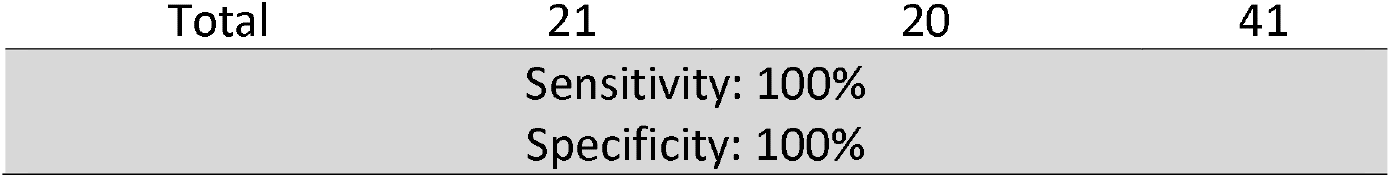
Sensitivity and specificity of HT-LAMP assay

## 4. Discussion

Based on RT-PCR, the current gold standard technique for detecting the SARS-CoV-2 virus in specimen swabs also needs laboratory-based viral extraction techniques [3]. There is an urgent need for alternatives that increase the possibilities for testing at the point-of-care setting and in limited settings, even if the present laboratory-based approach for SARS-CoV-2 is scalable and can be automated for high throughput. The current pandemic has, as expected, drawn attention to the use of LAMP for COVID-19 diagnostics. Although LAMP requires more work during the primer selection process, diagnosing it is much easier. LAMP has the benefit of having low sensitivity to impurities [12].

In this study, we confirm that one of the most rapid, straightforward, affordable, and scalable protocols for detecting SARS-CoV-2 nucleic acid is not only reliable but also highly specific when compared to RT-PCR detection. Most importantly, there is no RNA purification step in our HT-LAMP protocol, and this method does not need complicated equipment other than pocket bodywarmers and eye droppers. From sampling to detection, the protocol takes about 65 minutes, only requires a few reagents, and can even be self-performed by non-professionals. These characteristics enable the application of our methodology everywhere, even in a home setting.

In conclusion, our advancements over the current RT-LAMP procedures allow for the reliable and affordable detection of SARS-CoV-2. The findings serve as the foundation for further clinical performance investigations. We advise testing this protocol on a larger cohort of clinical specimens to further promote the use of HT-LAMP as a rapid home test for identifying COVID-19 patients. This SARS-CoV-2 detection technique can be used as a surveillance tool for sampling large populations after receiving additional validation.

Indeed, this method’s ease of use, product availability, and low cost will make it simple to monitor suspect infected individuals continuously. Additionally, this method can be applied in various places, such as workplaces, nursing homes, medical facilities, and points of entry. Last but not least, and equally crucial, this technique can be easily modified to check for infection by any other pathogen. If any new infectious pandemic emerges, all nations will need to have access to diagnostic testing for everyone in order to fight the pandemic more effectively. To satisfy this requirement, we anticipate that developing an RT-LAMP assay that is electro-equipment-free will be a significant advancement.

## Data Availability

All data produced in the present work are contained in the manuscript

## Ethics approval

The sample collection was approved by the hospital management. The internal use of samples was agreed upon under the medical and ethical rules of each participating individual. The study was approved by the Research Ethics Committee of Nguyen Tat Thanh University.

## Acknowledgements

The authors are also thankful to the staff of the local hospital for collecting samples and providing the epidemiological information.

## Conflicts of interest

The authors declare that they have no conflicts of interest.

## Notes

### Competing Interest Statement

The authors have declared no competing interest.

### Funding Statement

This study did not receive any funding

